# Tattoo practices and risk of hepatitis B and hepatitis C infection in the general population

**DOI:** 10.1101/2024.10.25.24316096

**Authors:** Milena Foerster, Marie Zins, Marcel Goldberg, Céline Ribet, Sofiane Kab, Bayan Hosseini, Rachel McCarty, Valerie McCormack, Khaled Ezzedine, Joachim Schüz

**Author notes:** Correspondence to: M Foerster. J Schüz. Contributorship statement: MF, KE and JS contributed to the planning of the work, MF, MZ, MG, CR, SK, and BH contributed to the conduct of the work and all authors contributed to the reporting of the work. MF is responsible for the overall content as guarantor and accepts full responsibility for the work and/or the conduct of the study, had access to the data, and controlled the decision to publish. The corresponding author attests that all listed authors meet authorship criteria and that no others meeting the criteria have been omitted. COI: All authors have completed the ICMJE uniform disclosure form at https://www.icmje.org/disclosure-of-interest/ and declare no conflict of interest. Copyright/Licence for publication: The Corresponding Author has the right to grant on behalf of all authors and does grant on behalf of all authors, a worldwide licence to the Publishers and its licensees in perpetuity, in all forms, formats and media (whether known now or created in the future), to i) publish, reproduce, distribute, display and store the Contribution, ii) translate the Contribution into other languages, create adaptations, reprints, include within collections and create summaries, extracts and/or, abstracts of the Contribution, iii) create any other derivative work(s) based on the Contribution, iv) to exploit all subsidiary rights in the Contribution, v) the inclusion of electronic links from the Contribution to third party material where-ever it may be located; and, vi) licence any third party to do any or all of the above.

## Abstract

**Objectives:** To prevent hepatitis B (HBV) and hepatitis C (HCV) infections and associated deaths from hepatocellular carcinoma and cirrhosis, better identification of transmission routes is needed. Here, we reassessed the impact of different tattooing practices on viral transmission.

**Design:** Population based cohort-study.

**Setting:** Cancer Risk Associated to the Body Art of Tattooing (CRABAT) cohort as part of the ongoing French national cohort study Constances (baseline examination from 2012-2018).

**Participants:** 110,402 participants (60,387 women and 50,015 men), of which 11.6% (12,789) were tattooed as per Constances follow-up questionnaire 2020. Complete exposure data on different exposure settings and countries of tattooing collected via complementary exposure assessment in 2023 (response rate 60%) was available for 7740 tattooed (4930 women and 2810 men) participants.

**Main outcome measures:** Self-reported HBV and HCV infections that were confirmed by surface antigen testing (HBsAG) and antibody (Anti-HCV) testing, respectively. Associations of different tattoo exposure characteristics (any tattoo; tattooed in/outside tattoo parlours; tattooed in/outside regulating countries; no tattoos (reference)) on subsequent HBV and/or HCV infections were assessed via multivariate logistic regression models, minimally adjusted and adjusted for known hepatitis risk factors, in the population >=45 years. Post-hoc, number of preventable HCV infections due to unsafe tattooing outside tattoo parlours was estimated.

**Results:** In fully adjusted models, tattooing was associated with increased risk of any hepatitis infection (Odds ratio (OR): 1.46 (95% confidence interval: 1.15; 1.86), with a particularly strong increased risk for HCV (2.26 (1.64; 3.11)) compared to HBV (1.08 (0.77; 1.52)) infection. The increased risk for HCV and to a lesser extend for HBV was due to tattooing outside tattoo parlours (HCV: 4.75 (2.81; 8.03); HBV: 1.88 (0.99; 3.57)) whereas tattooing outside regulating countries was associated with an increased risk for HCV (2.74 (1.00; 7.45) and HBV (1.96 (0.80; 4.84)). Risk of HBV and/or HCV were around 10-fold for tattooing outside tattoo parlours outside regulating countries. The estimated number of preventable HCV infections through safe tattoo practices was around 12,000 in France and over 150,000 in Europe.

**Conclusion:** The impact of unsafe tattooing practices as a preventable risk factor for HCV transmissions is highly underestimated.

**What is already known on the topic?:**

- Tattooing was identified as a potential transmission route for hepatitis infections in the early 1990s.
- Hygiene measures were implemented in tattoo parlours throughout (many) European countries to prevent bloodborne infection transmission through tattooing needles.
- Current hepatitis prevention strategies rarely/never consider tattooing as a common transmission route.

**What this study adds:**

- Unsafe tattooing practices are very common. One in four tattooed people got at least one tattoo outside parlours and one in five got tattooed in a country without strict hygiene regulations.
- Unsafe tattooing practices strongly increase the risk of HCV and to a lesser extend for HBV, making it the most important HCV transmission route after injecting drugs.
- The study provides evidence that raising awareness on unsafe tattooing and upscaling screening of persons that underwent unsafe tattooing might help to substantially reduce hepatitis infections and related morbidity and mortality.

## Introduction

It is estimated that worldwide 254 million people are living with Hepatitis B virus (HBV) infection and 50 million with Hepatitis C (HCV) virus infection. As a result of these infections, annually more than 1.3 million people are dying from liver cirrhosis and primary liver cancer.^1^ The World Health Organisation (WHO)’s global hepatitis strategy which was endorsed by all Member States, aims to reduce new infections by 90% and related deaths by 65% between 2016 and 2030.^2^ The International Agency for Research on Cancer (IARC) recommends HBV vaccination ideally in all new-borns and testing and treating for both HBV and HCV to help prevent liver cancer.^3^ Major known transmission routes are injecting drug use using shared needles for HCV, and vertical mother-child transmission for HBV, as well as several other transmission routes accounting for smaller numbers (e.g., sex between men, unscreened blood transfusions, or sharing of personal hygiene products).^2, 4, 5^ Consequently, current national and international prevention strategies target the respective risk populations. However, taken together only about 15% of HBV and 35% of HCV infections in Europe can be explained by these transmission routes underlining the need for the identification of further risk factors for hepatitis transmission and their respective high risk populations.^6, 7^

During the last two decades, tattooing increased as a common lifestyle trend with peak rates as high as 38% reporting at least one tattoo in the 35-40 age group in the UK.^8^ Tattooing could be a known yet widely underestimated risk factor of HBV/HCV infections and, due to its steeply rising worldwide popularity, even lead to an increase in those infections.^4, 9, 10^ Tattooing entails the needle-assisted injection of tattoo inks into the upper layer of the dermis, in most instances using a tattoo machine, naturally leading to blood contact. Consequently, tattoo-associated transmission of various blood-borne diseases is plausible and has been reported since decades.^11, 12^ A recent systematic review including 121 studies found a more than 200% and more than 50% elevated risk for HCV and HBV infections among tattooed individuals, respectively.^10^ However, the large majority of the included studies were purely descriptive and were not adjusted for other known transmission risk factors. Given the expected high correlation between tattooing and other risk-taking behaviours associated with hepatitis transmission, those risk estimates remain elusive.^13^

To better address this question, we use data from the “Cancer Risk Associated with the Body Art of Tattooing (CRABAT)” study to assess risk of HBV and HCV transmission following tattooing, adjusting for other transmission risk factors and stratified by different tattooing practices. We further extrapolate the number of HBV and HCV transmissions associated with different tattoo practices for the French and the European population, illustrating the immediate preventive potential.

## Methods

CRABAT is a prospective study of tattoo-associated long-term cancer risk nested in the French national cohort study called Constances.^14^ Constances consists of over 220,000 voluntary participants from the French metropolitan areas recruited to reflect the countries’ age, sex, and socioeconomic structure. It collects a wealth of self-reported data on sociodemographic and lifestyle factors as well as various medical and paramedical outcomes, starting from the baseline investigations between 2012 to 2018 and subsequent annual follow-up questionnaires. These data are enriched by record linkage to national health databases tracking all medical acts received and (fully or partialy) reimbursed by the national public health insurance in which all French citizens are automatically enrolled.

In 2020, the screening question (“Are you tattooed?“) was included in the Constances annual follow-up questionnaire to identify the tattooed study population within Constances for the CRABAT study. From July to December 2023, detailed tattoo exposure data were collected from the tattooed subgroup using a validated tattoo exposure questionnaire.^15^ In February 2024, the CRABAT study database was established merging the collected tattoo exposure data with sociodemographic, lifestyle and medical data for all participants having answered the 2020 follow-up questionnaire. The remaining untattooed Constances participants served as the unexposed comparison group.

### Tattoo exposure data

CRABAT collects detailed data on visual and contextual factors of tattooing using the validated exposure assessment tool EpiTAT, which was developed by piloting different methods of exposure assessment in a study comparing self-reported information with measured tattoos.^15^ For the present study, we extracted data on (i) total tattoo surface area in units of hand palms, (ii) the tattoo setting (5 multiple choice categories “By a professional in a tattoo parlour”, “By a professional in other circumstances, e.g. tattoo convention”, “By a lay person no matter where”, “Hand-poked or by traditional tattoo techniques not using a tattoo machine” (original categories combined into “Only in tattoo parlour”, “At least one tattoo outside parlour” to increase statistical power), (iii) the country where the tattoo was received from the question “Have you been tattooed outside France at least once? (yes/no)”, and if so, the respective countries in open text fields; tattoo countries were dichotomized into having at least one tattoo done outside European countries with tattoo hygiene regulations for at least 10 years (subsequently called “EU/EFTA countries with regulation”, the list of respective countries in supplementary List 1^16^) and other (iv) time period of tattooing (5 multiple choice categories “last year”, “>1 to <=5 years ago”, “>5 to <=10 years ago”, “>10 to <=15 years ago”, “15 years ago and longer”) to calculate the approximate date of tattoo acquisition by using the date of filling in the questionnaire minus the category midpoint of the earliest indicated category, and minus 20 years for the category “>15 years and longer”.

### Outcome data

Individual level HBV and HCV data were retrieved for the period 1^st^ January 2007-31^st^ December 2020 by record linkage to the national health database. This data included information on hospitalisation due to chronic HBV/HCV infection, as well as all conducted HBV/HCV tests during the respective years but not test results. We also retrieved self-reported data on infection status collected from baseline (“Have you ever been diagnosed with Hepatitis B? / Hepatitis C?”) or follow-up (“Since the last FUP, have you been infected with a liver disease?”) questionnaires between January 2012 and May 2021, included the respective age at diagnosis and the type of liver disease. We considered participants to have ever been infected with HCV or HBV (i) if they had been hospitalised for chronic infections; (ii) if a self-reported infection from the follow-up questionnaire was confirmed by at least two surface antigen tests for HBV (HBsAG) or two antibody tests (Anti-HCV) for HCV within the two years before or after the reported infection date; (iii) if a self-reported infection from baseline with infection date after 2007 was confirmed by HBsAG or Anti-HCV in the 2 years preceding or following the self-reported date; (iv) if a self-reported infection date before 2007 was confirmed by at least one HBsAG or Anti-HCV test after 2007. Participants who reported infection but without indication of testing after 2007 in the database or, for HBV, participants with only antibody testing (Anti-HBc or Anti-HBs) were not considered eligible for outcome analyses but are included in descriptive tables. Dates of infection were set to the date of hospital entry retrieved from national health data or to the corresponding self-reported infection date for participants who have not been hospitalised.

### Sociodemographic and lifestyle covariates

For the present analyses we extracted the following information from the Constances database: demographic variables including age, sex (gender not available in Constances), and highest educational level (according to the French education system); possible confounding variables including alcohol consumption (assessed using the Alcohol Use Disorders Identification Test (AUDIT) categorised to “Abstinent”, “Abuse”, “Dependence”, “Neither abuse nor dependence”, “Prefer not to answer”), lifetime number of sexual partners (categorised to “One”, “Two to five”, “Five to 10”, “10 and more”, “Prefer not to answer”), current condom use (“Yes”, “No”, “Not currently in a relationship”, and “Prefer not to answer”), sex of preferred sexual partner (“Male”, “Female” and “Prefer not to answer” to create the new variable called “Sex amongst men” with categories “No (women and heterosexual men)”, “Yes”, and “Prefer not to answer”). Injection drug use (IDU), known as the most important risk factor for HCV/HBV transmission during adulthood, was not assessed in Constances, therefore we used the national health data to identify participants having received opioid substitution therapy (in France either with butaprenorphine (median average dose 7.74mg/day) or with methadone (median average dose 43 mg/day)) at any time since 2009 and used this information as a proxy of ever IDU.^17^ HIV positive participants, known to belong to a population with higher HCV/HBV incidence, were also identified using the national health data.^5, 18, 19^

### Patient and public involvement

The EpiTAT questionnaire used in this study was developed in collaboration with tattoo artists and piloted with volunteers who were recruited via social media and printed flyers. The involvement of the public for developing this questionnaire informed the data collection methodology and analyses for this study. For the other aspects of this study, we did not directly involve patients or the public in the design or implementation of this research.

### Statistical analyses

Characteristics of tattoo exposures, sociodemographic and risk-profile variables were described overall, and separately for tattooed and non-tattooed participants and by HBV and HCV status. Potential differences across groups were assessed using the chi-squared test for categorical variables and the two-sided t-test for continuous variables. The relationships between tattooing and HBV or HCV infection status were analysed using multivariate logistic regression models using HBV and respectively HCV as dichotomous dependent variables; separate models were also calculated for HBV or HCV combined. the resulting odds ratio (OR) and 95% confidence intervals (CI) display the strength of association and related statistical uncertainty. Participants with infection dates preceding the first tattoo date were excluded from the analyses. As for participants with infection dates earlier than 1^st^ January 2007, timeliness of first tattoo and exposure could not be verified, therefore we assumed that the tattoo date proceeded the infection as first tattoos are usually acquired in early adulthood. Different models were fitted for (i) dichotomous exposure of being tattooed (yes/no), and (ii) by tattoo settings and tattoo country. For the tattooed population, a model including an interaction term for tattoo setting and tattoo country was fitted. Each model was fitted with two different levels of adjustment, a minimally adjusted model including sex and age as the only covariates and a fully adjusted model that additionally included educational level, alcohol use (AUDIT score), number of sexual partners, condom use, and sex amongst men. All analyses were restricted to persons who were 45 years or older in 2020 (i.e. the year of the initial tattoo assessment), as few HBV and HCV cases could be identified in younger age-groups. We also excluded participants who received opioid replacement therapy (0.1% of the total population; 0.4% amongst the tattooed participants) and HIV positive persons (0.8%; 0.6%) from the main analyses given their small numbers but strong association with the outcome to not dilute the analysis of associations with the exposure of interest; however, we looked at them separately.

Sensitivity analyses were fitted exploring the relationship between tattoo exposure variables and HBV/HCV infections (i) for the tested or screened population including only persons with known HBV/HCV testing recorded in the national health database (ii) for a low-risk group excluding both men who had sex with men, participants with >10 sexual partners, and participants with alcohol abuse (iii) by sex; and (iv) for the binary association with tattooing to responders vs non-responders of our exposure questionnaire; (v) We also restricted the analyses to infections with known timeliness of tattoo and infection date (participants diagnosed after 31^st^ December 2006). Finally, risk for tattooed vs non tattooed was calculated for the high-risk groups of ever injecting drug users or HIV positive participants (due to limited sample sizes these were only adjusted for age, sex, education, and either HIV or ever injected drug use).

Finally, we estimated the number of HBV and HCV infections due to unsafe tattoo practices (i.e., tattooing outside tattoo parlour) in the French and European population for ages 45 to 75 years. The tattooed population was estimated by extrapolating the Constances prevalence of unsafe tattoo practices (defined as having received at least one tattoo outside tattoo parlours for HCV and being tattooed outside regulating countries for HBV) to UN data, applying separate rates per 10 year-age bands. To calculate the number of prevalent HBV/HCV infections we applied the Constances population prevalence in ages 45 and older to the UN data. The number of attributable and therefore preventable cases were then calculated by multiplying the population prevalence for HBV/HCV with the number of persons that underwent unsafe tattoo practices and with the risk estimate observed in our study for people who underwent unsafe tattoo practices. This was done for the population of France and for the European population with Europe defined by the 2018 revised version of the United Nations World Urbanization Prospects.^20^ The resulting numbers reflect the preventable HBV/HCV infections in the respective populations of 45-75 years in 2022, with these numbers increasing with each younger age group entering this age interval over time.

## Results

Of all 110,402 eligible participants, 11.6% (n=12,789) reported to have at least one tattoo (supplementary Figure 1). Tattoo prevalence was higher in women compared to men (Table 1) and steeply decreased from the youngest age group of under 35 years (23.8%, n=2,746/11,520) to the oldest >65 (2.8%, n=770/27,050; data not shown) resulting in an inversed distribution of tattooing and age distribution (Table 2). There was a tendency for tattooed individuals to have lower educational levels on average. Alcohol abuse was higher in tattooed persons as well as the number of sexual partners, engagement in sex amongst men and current condom use. Of the study sample 823 (0.8%) participants had ever been infected with HBV and 476 (0.4%) with HCV, with higher proportions amongst tattooed compared to non-tattooed participants for both infections. Individuals with hepatitis infections showed a similar sociodemographic and lifestyle risk profile than the tattooed subgroup (Supplementary Table 1). Only about one third of overall participants were tested for HBV or HCV, contrasted by about half of the tattooed population (Table 2).

**Table 1:** Exposure and Outcome distributions in the study population by sex, and by infection status.

|  | Men<br>50,015 | Women<br>60,387 | HBV<br>823 (0.8%) | HCV<br>476 (0.4%) | Overall<br>110,402 |
| --- | --- | --- | --- | --- | --- |
| Tattooed | 4774 (9.5 %) | 8015 (13.3 %) | 77 (9.4 %) | 88 (18.5 %) | 12789 (11.6 %) |
| Responding to EpiTAT | 2810 (5.6 %) | 4930 (8.2 %) | 59 (7.2 %) | 41 (8.6 %) | 7740 (7 %) |
| **Setting<sup>A</sup>** |  |  | *Of these:* |  |  |
| Only in parlour | 2078 (74.6 %) | 4064 (83.4 %) | 28 (62.2 %) | 17 (41.5 %) | 6142 (80.2 %) |
| At least one tattoo outside parlour | 709 (25.4 %) | 809 (16.6 %) | 17 (37.8 %) | 24 (58.5 %) | 1518 (19.8 %) |
| **Place/country<sup>B</sup>** |  |  |  |  |  |
| EU/EFTA countries with regulation | 2448 (87.7 %) | 4346 (89 %) | 36 (80 %) | 35 (85.4 %) | 6794 (88.5 %) |
| No or recent regulation | 345 (12.3 %) | 540 (11 %) | 9 (20 %) | 6 (14.6 %) | 885 (11.5 %) |

<sup>A</sup> 83 missing values in tattoo context<sup>B</sup> 64 missing values in tattoo country

**Table 2:** Demographic characteristics and risk profile, overall and by tattoo status.

|  | Overall<br>110,402 | Not tattooed<br>97,613 | Tattooed<br>12,789 | p-value |
| --- | --- | --- | --- | --- |
| **HBV** | 823 (0.7 %) | 746 (0.8 %) | 77 (0.6 %) | 0.045 |
| **Tested for HBV** | 39622 (35.0 %) | 33267 (33.2 %) | 6355 (48.4 %) | <0.001 |
| **HCV** | 476 (0.4 %) | 388 (0.4 %) | 88 (0.7 %) | <0.001 |
| **Tested for HCV** | 42712 (37.7 %) | 35871 (35.8 %) | 6841 (52.1 %) | <0.001 |
| **Sex** |  |  |  |  |
| Male | 50015 (45.3 %) | 45241 (46.3 %) | 4774 (37.3 %) | <0.001 |
| Female | 60387 (54.7 %) | 52372 (53.7 %) | 8015 (62.7 %) |  |
| **Age** |  |  |  |  |
| <=35 | 11520 (10.4 %) | 8774 (9.0 %) | 2746 (21.5 %) | <0.001 |
| >35-<=45 | 21310 (19.3 %) | 17482 (17.9 %) | 3828 (29.9 %) |  |
| >45-<=55 | 24489 (22.2 %) | 21083 (21.6 %) | 3406 (26.6 %) |  |
| >55-<=65 | 25263 (22.9 %) | 23224 (23.8 %) | 2039 (15.9 %) |  |
| >65 | 27820 (25.2 %) | 27050 (27.7 %) | 770 (6.0 %) |  |
| **Education** |  |  |  |  |
| No diploma | 1789 (1.6 %) | 1443 (1.5 %) | 346 (2.7 %) | <0.001 |
| General education certificate | 5248 (4.8 %) | 4654 (4.8 %) | 594 (4.6 %) |  |
| Vocational training certificate | 14973 (13.6 %) | 12784 (13.1 %) | 2189 (17.1 %) |  |
| Baccalaureate or equivalent diploma | 16600 (15.0 %) | 13941 (14.3 %) | 2659 (20.8 %) |  |
| Baccalaureate + 2 or 3 years | 29552 (26.8 %) | 25723 (26.4 %) | 3829 (29.9 %) |  |
| Baccalaureate + 4 years | 10272 (9.3 %) | 9428 (9.7 %) | 844 (6.6 %) |  |
| Baccalaureate + 5 and more | 29968 (27.1 %) | 27849 (28.5 %) | 2119 (16.6 %) |  |
| Missing | 2000 (1.8 %) | 1791 (1.8 %) | 209 (1.6 %) |  |
| **Alc (audit score)** |  |  |  |  |
| Abstinent | 3273 (3.0 %) | 3014 (3.1 %) | 259 (2.0 %) | <0.001 |
| Abuse | 17148 (15.5 %) | 14563 (14.9 %) | 2585 (20.2 %) |  |
| Dependent | 4258 (3.9 %) | 3480 (3.6 %) | 778 (6.1 %) |  |
| Neither Abuse nor debendence | 79002 (71.6 %) | 70692 (72.4 %) | 8310 (65.0 %) |  |
| Missing | 6310 (5.7 %) | 5509 (5.6 %) | 801 (6.3 %) |  |
| **Ever opioid replacement therapy** | 146 (0.1 %) | 90 (0.1 %) | 56 (0.4 %) |  |
| **HIV positive** | 823 (0.7 %) | 746 (0.8 %) | 77 (0.6 %) | 0.045 |
| **Number of sexual partners** |  |  |  |  |
| 1 partner | 17516 (15.9 %) | 16410 (16.8 %) | 1106 (8.6 %) | <0.001 |
| 2-5 partners | 30968 (28.1 %) | 27522 (28.2 %) | 3446 (26.9 %) |  |
| 6-10 partners | 16247 (14.7 %) | 13739 (14.1 %) | 2508 (19.6 %) |  |
| >10 partners | 11041 (10.0 %) | 8937 (9.2 %) | 2104 (16.5 %) |  |
| Prefer not to answer | 23178 (21.0 %) | 20561 (21.1 %) | 2617 (20.5 %) |  |
| Missing | 11452 (10.4 %) | 10444 (10.7 %) | 1008 (7.9 %) |  |
| **Sex amongst men** |  |  |  |  |
| Women or never | 45622 (91.2 %) | 41403 (91.5 %) | 4219 (88.4 %) | <0.001 |
| Ever | 3198 (6.4 %) | 2730 (6.0 %) | 468 (9.8 %) |  |
| Prefer not to answer | 1195 (2.4 %) | 1108 (2.5 %) | 87 (1.8 %) |  |
| **Current condom use** |  |  |  |  |
| Yes | 27607 (25.0 %) | 23521 (24.1 %) | 4086 (31.9 %) | <0.001 |
| No | 59526 (53.9 %) | 52937 (54.2 %) | 6589 (51.5 %) |  |
| Not currently in a relationship | 8846 (8.0 %) | 7738 (7.9 %) | 1108 (8.7 %) |  |
| Missing | 14423 (13.1 %) | 13417 (13.7 %) | 1006 (7.9 %) |  |

For all 7,740 eligible tattooed participants answering the EpiTAT questionnaire on details of the tattoo exposure in 2023 (total respondents 7,928; response rate 60.4%), and with available outcome data, the median tattoo surface size was 1 hand palm (Inter-quartile range (IQR) 0.5; 3) and almost half of all participants got their first tattoo 15 years ago or longer (46.3%, n=3,555/7674, data not shown). The large majority of participants received their tattoo(s) exclusively in tattoo parlours, but one in five persons had at least one tattoo from outside a tattoo parlour and over one in 10 tattooed persons reported at least one tattoo from outside EU/EFTA regulating countries (Table 1).

Tattooed participants had a 1.6-fold elevated risk of either hepatitis infections in the minimally adjusted models, slightly attenuated to 1.5 through full adjustment (Table 3). However, when analysed by tattoo context, the risk was confined to those who had at least one tattoo outside of tattoo parlours and/or outside EU/EFTA regulating countries culminating in an almost 10-fold increase for those tattooed outside parlours outside EU/EFTA regulating countries. A decreased risk for any hepatitis infection was seen for participants who got tattooed inside tattoo parlours.

**Table 3:** Odd’s ratios and 95% CIs of the multivariate logistic regression models on risk of chronic Hepatitis B or chronic Hepatitis C virus infection and tattoo exposure variables for two different levels of adjustment.

|  | HBV or HCV |  |  |  | HBV |  |  |  | HCV |  |  |  |
| --- | --- | --- | --- | --- | --- | --- | --- | --- | --- | --- | --- | --- |
|  | Model 1<sup>A</sup> |  | Model 2<sup>B</sup> |  | Model 1<sup>A</sup> |  | Model 2<sup>B</sup> |  | Model 1<sup>A</sup> |  | Model 2<sup>B</sup> |  |
|  | OR | 95% CI | OR | 95% CI | OR | 95% CI | OR | 95% CI | OR | 95% CI | OR | 95% CI |
| **Tattooed** | 1.58 | (1.25; 1.99) | 1.46 | (1.15; 1.86) | 1.11 | (0.80; 1.54) | 1.08 | (0.77; 1.52) | 2.69 | (1.98; 3.66) | 2.26 | (1.64; 3.11) |
| **Tattoo context** |  |  |  |  |  |  |  |  |  |  |  |  |
| not tattooed |  | ref |  | ref |  | Ref |  | Ref |  | ref |  | ref |
| only in tattoo parlour | 0.55 | (0.31; 0.95) | 0.48 | (0.26; 0.88) | 0.50 | (0.25; 1.01) | 0.43 | (0.20; 0.91) | 0.69 | (0.31; 1.56) | 0.63 | (0.28; 1.42) |
| outside tattoo parlour | 2.74 | (1.78; 4.23) | 2.42 | (1.49; 3.92) | 1.80 | (0.95; 3.38) | 1.88 | (0.99; 3.57) | 5.54 | (3.31; 9.25) | 4.75 | (2.81; 8.03) |
| **Tattoo country** |  |  |  |  |  |  |  |  |  |  |  |  |
| Not tattooed |  | ref |  | ref |  | Ref |  | Ref |  | ref |  | ref |
| EU/EFTA countries with regulation | 0.98 | (0.67; 1.43) | 0.93 | (0.63; 1.37) | 0.67 | (0.39; 1.17) | 0.62 | (0.35; 1.12) | 1.74 | (1.07; 2.82) | 1.55 | (0.95; 2.53) |
| No or recent regulation | 2.07 | (0.97; 4.40) | 1.85 | (0.86; 3.96) | 2.17 | (0.89; 5.28) | 1.96 | (0.80; 4.84) | 3.24 | (1.20; 8.76) | 2.74 | (1.00; 7.45) |
| **Interaction amongst the tattooed** |  |  |  |  |  |  |  |  |  |  |  |  |
| In studio in regulating country |  | ref |  | ref |  | Ref |  | Ref |  | Ref |  | Ref |
| Outside parlour in regulating country | 4.65 | (2.04; 10.58) | 6.44 | (2.68; 15.48) | 2.58 | (0.80; 8.30) | 4.00 | (1.12; 14.30) | 7.59 | (2.58; 22.33) | 7.76 | (2.59; 23.30) |
| Inside parlour outside in regulating country | 2.19 | (0.60; 8.03) | 2.27 | (0.59; 8.78) | 2.34 | (0.47; 11.68) | 2.54 | (0.45; 14.21) | 1.48 | (0.17; 12.71) | 1.45 | (0.16; 12.84) |
| Outside parlour outside in regulating country | 8.70 | (3.05; 24.83) | 9.85 | (3.22; 30.12) | 8.83 | (2.39; 32.58) | 13.26 | (3.13; 56.15) | 12.14 | (3.14; 46.84) | 10.97 | (2.70; 44.50) |

<sup>A</sup>Adjusted for sex, age at initial tattoo assessment 2020
<sup>B</sup>Adjusted for sex, age at initial tattoo assessment, highest educational level, alcohol consumption (audit score), number of sexual partners, condom use, and sex amongst men

Looking at hepatitis subtypes separately, no risk difference was seen in tattooed compared to non-tattooed individuals for HBV. However, risk was about 2-fold elevated for being tattooed outside a tattoo parlour as well as outside EU/EFTA regulating countries though the estimates failed to reach statistical significance. Amongst the tattooed population, though numbers were small, the interaction of tattoo setting and tattoo country showed a strong association with a 13-fold increased risk for participants tattooed outside parlours outside EU/EFTA regulating countries. As for any infection. a decreased risk for HBV infection was seen for participants who got tattooed inside tattoo parlours reaching statistical significance in the fully adjusted model.

For HCV, tattooed individuals had an about 2.7-fold higher risk of HCV infection, attenuated to 2.3-fold with full adjustment. Regarding different tattoo settings, HCV risk was strongly elevated for individuals being tattooed outside tattoo parlours; the risk remained 4.8-fold after full adjustment and was also 2.8-fold increased among those tattooed outside EU/EFTA regulating countries. Again, risk was decreased for participants who got tattooed inside studio though not reaching statistical significance. The subgroup analyses of the interaction of tattoo setting and tattoo country in the tattooed population only, revealed again an 11-fold increased risk for individuals that have been tattooed outside tattoo parlours outside European regulating countries, with a lower confidence limit of a more than 2.7-fold effect, although numbers became small. Compared to other adjustment factors, tattoo context and tattoo country showed generally the strongest effects (Supplementary Figures 2 and 3).

Sensitivity analyses showed, despite lack of power, similar results to main analyses (Supplementary Tables 2-4). When restricted to the tested population only, Figures 1 and 2 show slightly attenuated estimates. However, for this subgroup, estimates for the known risk factors decreased considerably compared to the main analyses pointing towards selection bias in this group (supplementary Figures 4 and 5). For the low-risk group as defined in the Methods, there was a tendency for stronger effects for HBV but weaker effects for HCV. Stratification by sex showed stronger estimates for being tattooed overall for men compared to women whereas the analysis of different tattoo circumstances seemed entirely driven by women for HBV, and to a lesser extend for tattoo context and HCV. Restricting the analyses to participants with confirmed time of tattoo exposure proceeding infection, possible for more recent infections in participants diagnosed after 2007, confirmed the main association with an over two-fold increased risk for tattooed vs non-tattooed participants, and a 6.4-fold elevated risk for those with at least one tattoo received outside parlours (Supplementary table 4).

**Figure 1:**
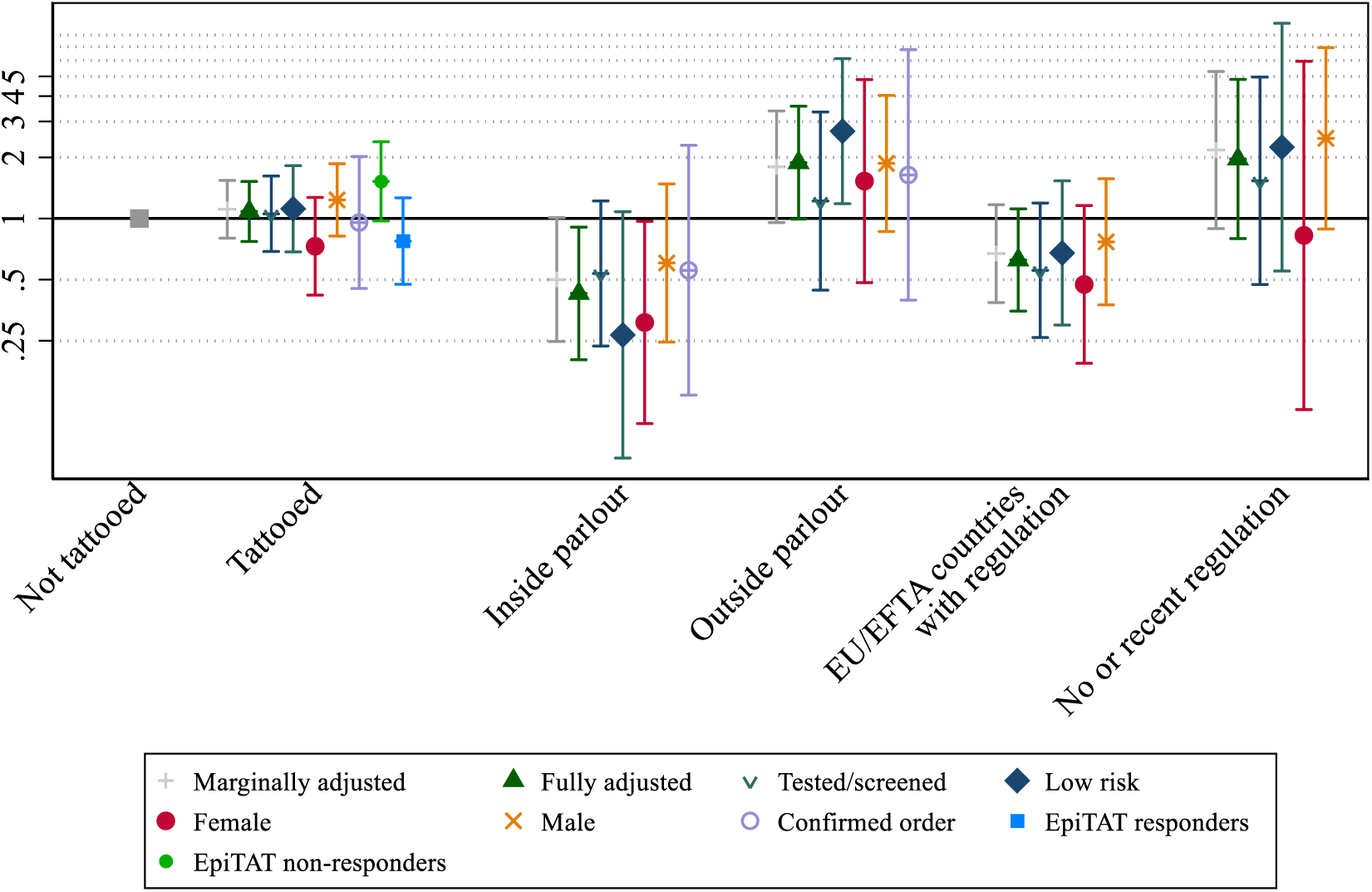
Odd’s ratios for tattoo associated Hepatitis B infection by tattoo setting, for basic and fully adjusted models and sensitivity analyses in the tested group, a low risk group, by biological sex, for different ways to calculate date of first tattoo. Due to small cell numbers in the tattoo context variable in “EU/EFTA counties with regulation” vs “No or recent regulation”, sensitivity analyses could not be calculated for HBV infections with confirmed tattoo-infection order (cases after 31^st^ december 2006).

**Figure 2:**
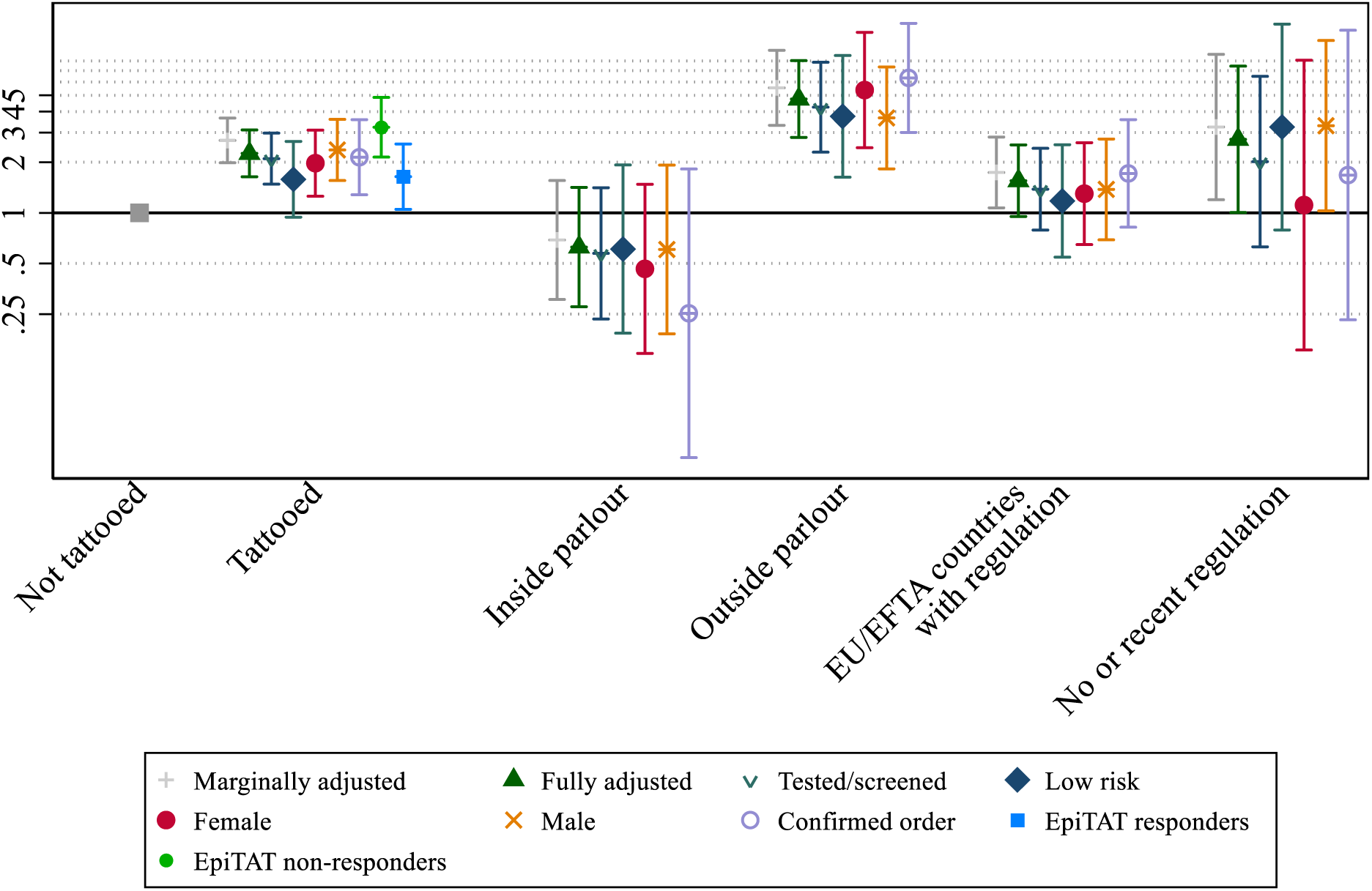
Odd’s ratios for tattoo associated Hepatitis C infection by tattoo setting, for basic and fully adjusted models and sensitivity analyses in a low risk group, by biological sex, for different ways to calculate date of first tattoo.

Exploratory analyses found both ever having had an opioid replacement therapy (with an OR of 6.69 (CI: 2.53; 17.70) for HBV and of 22.54 (CI: 12.24; 41.50) for HCV), as well as being HIV positive (with an OR of 5.12 (CI: 3.21; 8.15) for HBV and of 10.10 (CI: 5.90-17.31) for HCV) strongly related to the hepatitis infections, confirms our rationale for excluding them from the main analysis on the association between tattooing and the infections (data not shown).

The estimation of HCV cases attributable to unsafe tattoo practices defined as having at least one tattoo outside a tattoo parlour in the population of 45-75 years in France, assuming the infection and tattoo prevalence proportions from Constances, revealed 11,969 preventable HCV transmissions. Extrapolating to the European scale the numbers of preventable infections due to unsafe tattoo practices would increase to 145,374 cases of HCV. These numbers increase with each age group entering the 45-75 years group over time. We did not estimate attributable cases for HBV because of the less robust estimates. Furthermore, through the roll-out of HBV vaccination in most European countries and globally, we expect the tattoo-related HBV infections to decline strongly in the upcoming years.

## Discussion

In this study, we found a more than two-fold increased risk of HCV infection in tattooed individuals compared with non-tattooed individuals overall, whilst risk for HBV infection was only slightly increased. While the good news is that this was not true for tattoos done in tattoo parlours, we found a strong increase in risk for HCV and to a much lesser extent HBV associated with tattooing outside of tattoo parlours and also outside of European countries with strict hygiene regulations. The risks culminated in a more than 10-fold increase in HBV and/or HCV infection in people with tattoos done outside of a tattoo parlour and outside of these countries with regulations. This leads to an estimate of about 15,000 avoidable HBV/HCV infections in France, extrapolated to almost 145,000 for Europe, in the 45-75 age group. When we restricted analyses to 2007 or later, to ensure tattoos were received prior to infection, receiving a tattoo outside of a tattoo parlour was associated with an over 6-fold increased risk of HCV, even higher than in the main analyses. As we observed an even stronger risk for more recent infections, this suggests risk of HCV from unsafe tattooing practices is a concern for younger generations. This underscores the high likelihood of unsafe tattooing as an underestimated risk factor for HCV/HBV transmission, and from our data would make unsafe tattooing the second most important risk factor after injecting drug use for HCV.

To our knowledge, this is the first large epidemiological study to estimate the risk of HCV and HBV infection associated with different tattooing practices, which is a public health priority given the already high and increasing proportion of tattooed individuals worldwide and the recent rebound of new HCV infections in many parts of the world.^21^ (Notably, the UK official incidence estimates decline but their estimation is based on the number of injecting drug users and roll-out of treatment and prevention strategies in that group alone.^22^) Strengths of our study are, the rich data source using a nationwide prospective cohort study and the detailed tattoo exposure data using a validated exposure assessment instrument. Results for known transmission pathways other than tattooing are as expected, confirming the generalisability of our results. But there are also limitations. First, the analyses were cross-sectional with retrospective exposure and outcome assessment and reported estimates can only be interpreted as adjusted prevalence ratios of infection. Outcome ascertainment relied mostly on a combination of self-reported data and test dates from national health insurance data. While the latter is an objective source, for confidentiality reasons it may not provide test results. Consequently, some outcome misclassification cannot be ruled out. However, limiting the analyses to the tested population yielded similar results to the main analyses which is reassuring. Further, in contrast to the general population, HBV/HCV incidence is highest in age groups under 45 years, yet only few HBV/HCV cases were identified in the Constances population in this age group. While this probably owes to the cohort’s design, it forced us to restrict the analyses to the older age groups. We cannot rule out that risk estimates might be slightly attenuated in younger age groups if we assume that today’s conditions outside tattoo parlours benefitted from improvements of hygiene standards inside tattoo parlours; however, it seems more likely that lower HBV/HCV rates in younger age groups in our study are symptomatic for a healthy cohort effect and/or underdiagnosis, due to under-presentation of individuals that are at high risk and routinely screened in a general population cohort with voluntary participation. Consequently, only one third of study participants having ever got tested for HBV/HCV. As in that population tattooing was more common (16% vs 9%, data nor shown) we deliberately choose to calculate our main analysis on the whole sample to avoid biased estimates. The lower estimates for the tattoo circumstances and the strong attenuation for known risk behaviours such as sex amongst men or multiple sexual partners when limiting analysis to the screened population demonstrate that bias. Furthermore, the finding of reduced infection risk when tattooed inside tattoo parlours was surprising. While we tried further analyses (age-stratification, exclusion of participants who got tattoos in countries with higher HBV infection rates), the risk reduction remained stable. As we do not have another explanation for this finding, we cannot rule out that due to our retrospective design some exposure misclassification took place (e.g. participants with infection might have made them more likely to report tattoos outside parlours).

Our estimates for binary exposure and risk of infection are in line with recent literature.^10^ To our knowledge the relationship between unsafe tattoo practices and hepatitis infections has only been investigated in two studies dating back 15 years and longer. The respective studies on risk factors for HCV transmission examined relations with unsafe tattooing and found similar estimates but samples were small and homogenous (5,000 college students in Texas and 200 healthy young men in Taiwan) hampering generalisation. ^23, 24^ After identification of tattooing as a risk factor for bloodborne disease transmission in the 1990s, many industrialised countries implemented hygiene standards for tattoo parlours (in Europe e.g. DIN EN 17169:2020) in which professional tattoo artists today practice under sterile conditions.^10,11^ However, in our tattooed population embedded in a large-scale nationwide cohort in France, one in five tattooed persons reported at least one tattoo from outside a tattoo parlour. This corresponds to a large population at risk considering today’s tattoo prevalence of up to 30-40% in age groups under 40 years in industrialised countries^8^. As unsafe tattoo practices were never studied with sufficient detail on a population-level, its risk might have been underestimated. Moreover, tattooing has become more and more popular globally. Tattoo prevalence estimates of 22% in Brazil, and 12% in China and Russia indicate popularity around the world.^25^ In this context, our finding of an increased risk for HBV/HCV infections if tattooed outside regulating countries which is particularly strong if not tattooed in a tattoo parlour, emphasize the urgent need to scale-up guidelines for safe tattooing practices on a global level. Given that unsafe tattooing was amongst the strongest risk factors in our study, medical personal should be better aware on tattoo-associated transmission risks and recommend testing to respective patients in order to prevent a global tattoo-associated increase of HBV/HCV infections.

Our results further emphasize the need to scale-up HBV vaccination to immunize newborns, children and adolescents up to age 15 years, the latter age group even more important considering that today many people get a first tattoo in early adulthood. While coverage reaches up to 95% in the UK, vaccination is still not mandatory in many European countries.

Finally, the results call for future research. Tattoo exposure data on a global scale is urgently needed to address associated risk of bloodborne virus infections, particularly in endemic areas. Also, recent research found an increased risk of lymphoma related to tattoos.^26^ In that study, the proposed biologic mechanisms of this association was the potential toxicity of tattoo pigments and their accumulation in the lymph nodes (also the reason why tattoo inks have recently been included in a priority list for carcinogenic evaluation by the advisory group for the Monograph programme of the IARC).^27, 28^ However, as HCV infections like microbial infections, both transmitted by unsafe tattooing itself are strong risk factor for lymphoma, the present results underscore the need to examine the role of confounding through tattoo-associated infections in estimating cancer risks from tattoo inks.^29–31^

### Ethics approval

The Constances study was approved by the Institutional review board (IRB) of the French Institute of Health (Inserm) (Opinion n°01-011, then n°21-842), and authorized by the by the French Data Protection Authority (“Commission Nationale de l’Informatique et des Libertés”, CNIL) (Authorization #910486). The CRABAT study received additional approval by the IARC Ethics Committee (IEC 22-02), and was authorized by the CNIL (#22015584).

### Data sharing statement

For data security reasons to protect participants, the data of the present analysis is hosted on the secured data access platform CASD (“Centre d’Accès Sécurisé aux Données”) whose access, via a dedicated SD-box, is strictly regulated. Consequently, individual level data cannot be shared. All data access requests for other research purposes needs to be formally addressed to the Constances and CRABAT study teams via the application procedures outlined on the Constances website (www.constances.fr).

## Data Availability

All data produced in the present study are available upon reasonable request to the authors and the Constances cohort.

## Acknowledgements

The authors thank the team of the “Population-based cohorts unit” (Cohortes en population) that designed and manages the Constances cohort study. They also thank the French national health insurance fund (“Caisse nationale d’assurance maladie”, Cnam) and its Health screening centres (“Centres d’examens de santé”), which are collecting a large part of the data, as well as the French national old-age insurance fund (“Caisse nationale d’assurance vieillesse”, Cnav) for its contribution to the constitution of the cohort, and ClinSearch, Asqualab and Eurocell, which are conducting the data quality control. Special Thanks go to Prof Francesco Negro, senior hepatologist from Turin, Italy, for his helpful counselling in hepatology.

## Disclaimer

Where authors are identified as personnel of the International Agency for Research on Cancer/ or the World Health Organization, the authors alone are responsible for the views expressed in this article and the views do not necessarily represent the decisions, policy, or views of the International Agency for Research on Cancer/ or the World Health Organization.

