## Supplemental material for "Tattoo practices and risk of hepatitis B and hepatitis C infection in the general population"

### Supplementary material

#### Supplementary List

List of European countries with hygiene regulations for longer than 10 years as summarised in *Piccinini P, Bianchi I, Pakalin S, Senaldi C. Safety of tattoos and permanent make-up Compilation of information on legislative framework and analytical methods. EUR27394. 2015.*

The recommendations of the Czech Republic and Finland were not considered due to their vagueness. The list below always cites the first meaningful national document.

Austria: Regulation of the Minister of Economy and Labour on rules of exercise for pedicure, cosmetics and massage professionals, 2008

Belgium Arrêté Royal (A.R.) 25/11/2005 réglementant les tatouages et les piercings, 2005

Denmark: Recommendation from the Danish EPA on the safety of tattoo inks, 2014

France: Order 2008-149 of 19 February 2008 establishing hygiene requirements for tattooing and piercing processes, modifying the public health code

Germany: Ordinance [of the Federal Länder] on the prevention of communicable diseases in certain professional activities, since the 90ies

Italy: Order of the Health Ministry 05/02/1998 no. 2·9/156 - Guidelines for the implementation of safety procedures for tattooing and piercing, 1998

Malta: Tattoo Studios and Tattooing (Conditions) Regulations, 2011

Romania: Order n° 1136/2007 on the hygiene standards for the cabinets of body beauty, (OJ of Romania n°484/2007)

Slovakia: Ordinance of Ministry of Health No. 554/2007 Coll. on requirements on facilities of human body care

Slovenia: Rules on minimum sanitary and health requirements for hygiene care and other similar establishments, 2009

Spain: Inter-regional Council of the National Health System 2003 and regulations of autonomous regions, establishing the hygienic requirements of tattoo and piercing parlours, 2003

Sweden: Ordinance (AFS 2005:1) on microbiological work risks, contamination, toxic effects and sensibilisation (Swedish Work Environment Authority), 2005

The Netherlands: Ministry Regulation on the use of tattooing/piercing materials (23·05·07, n° VGP/PSL 2770998), 2007

United Kingdom: "Tattooing and body piercing guidance toolkit", issued by the Health and Safety Laboratory (HSE) in 2013.

Supplementary Figure 1: Flow Chart of study sample

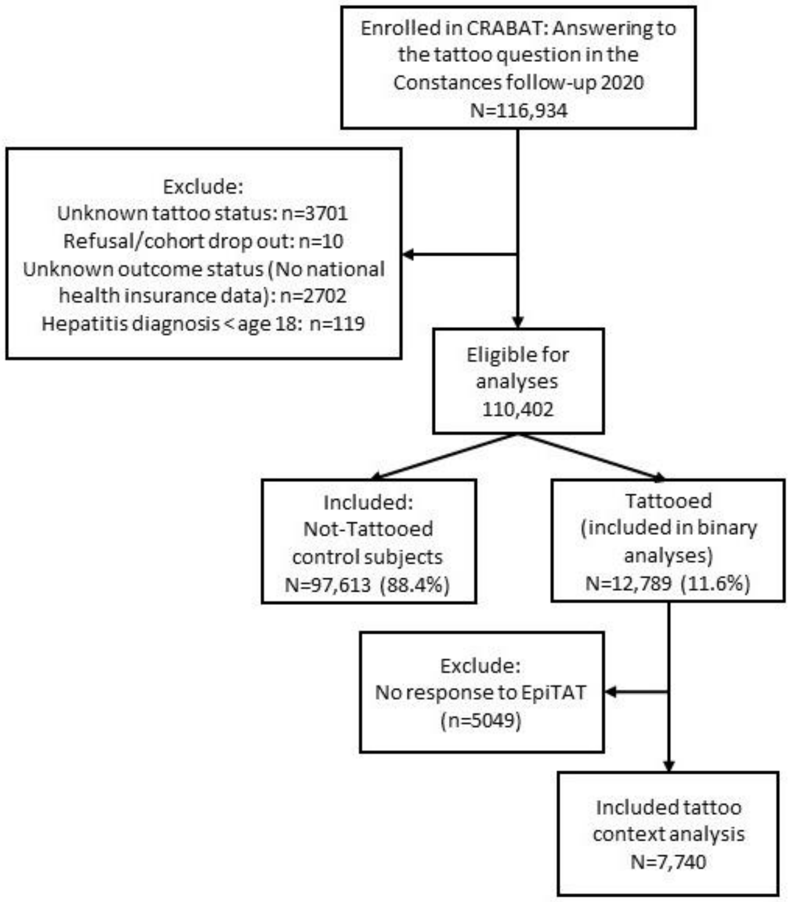

Supplementary Figure 2: ORs for tattoo exposure variables in different sensitivity analyses for all hepatitis infections.

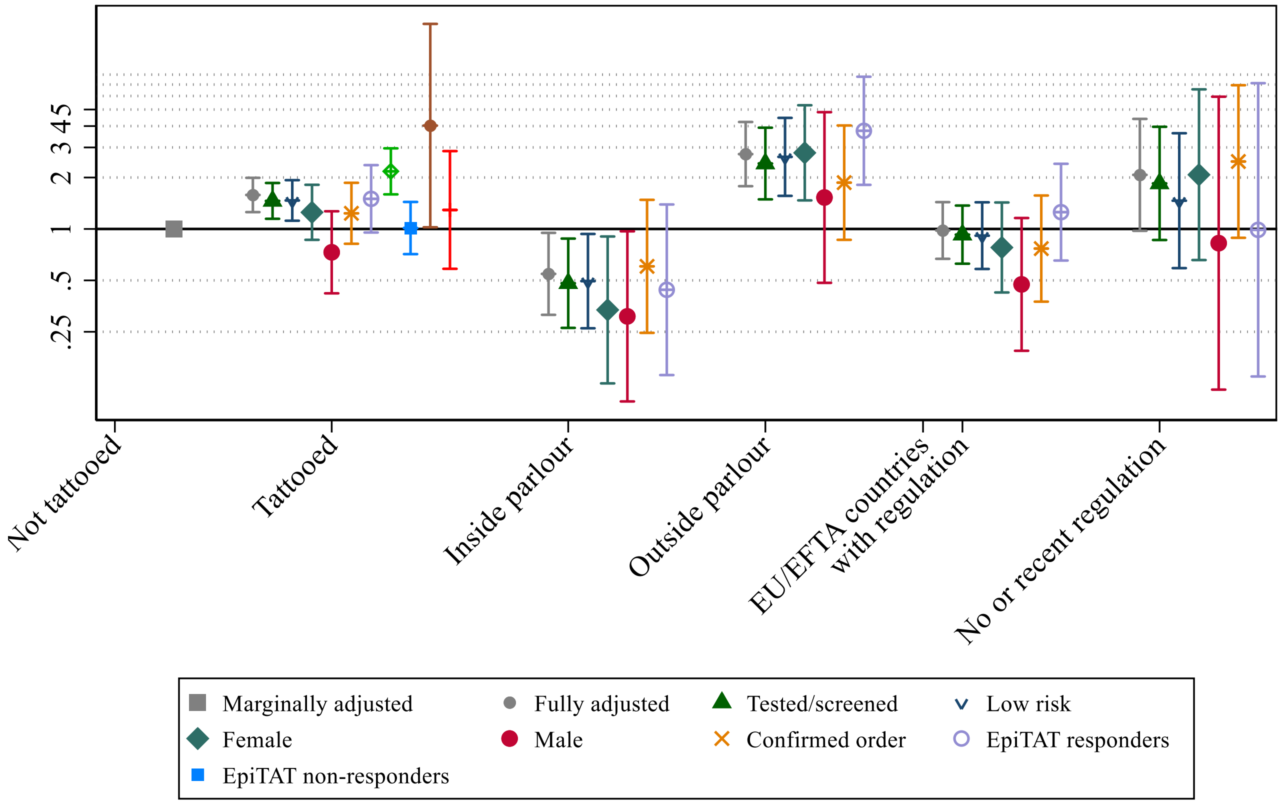

Supplementary Figure 2: ORs for Hepatitis B infection for covariates included in the fully adjusted models (adjusted for tattooed (yes/no)).

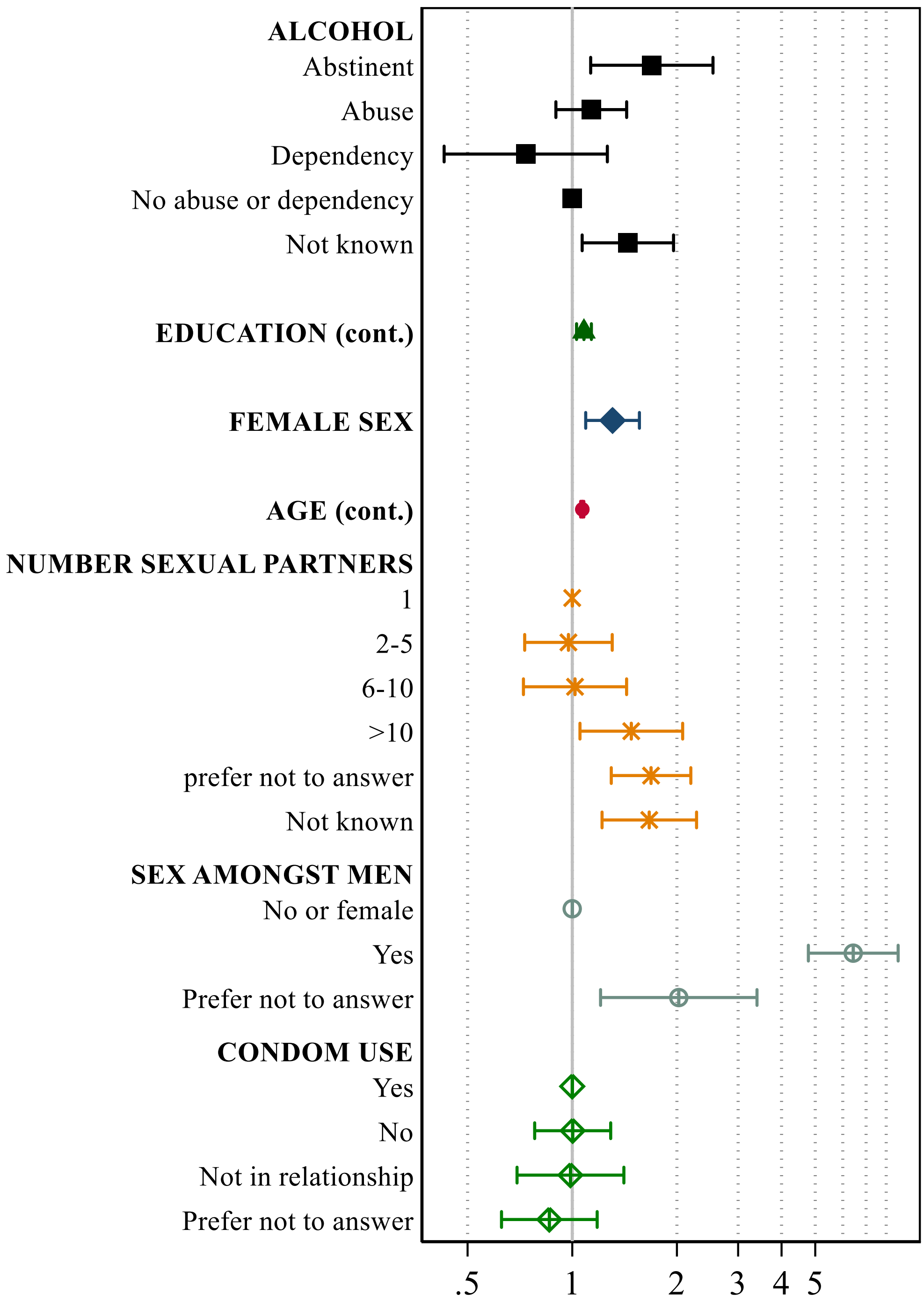

Supplementary Figure 3: ORs for Hepatitis C infection for covariates included in the fully adjusted models (adjusted for tattooed (yes/no))

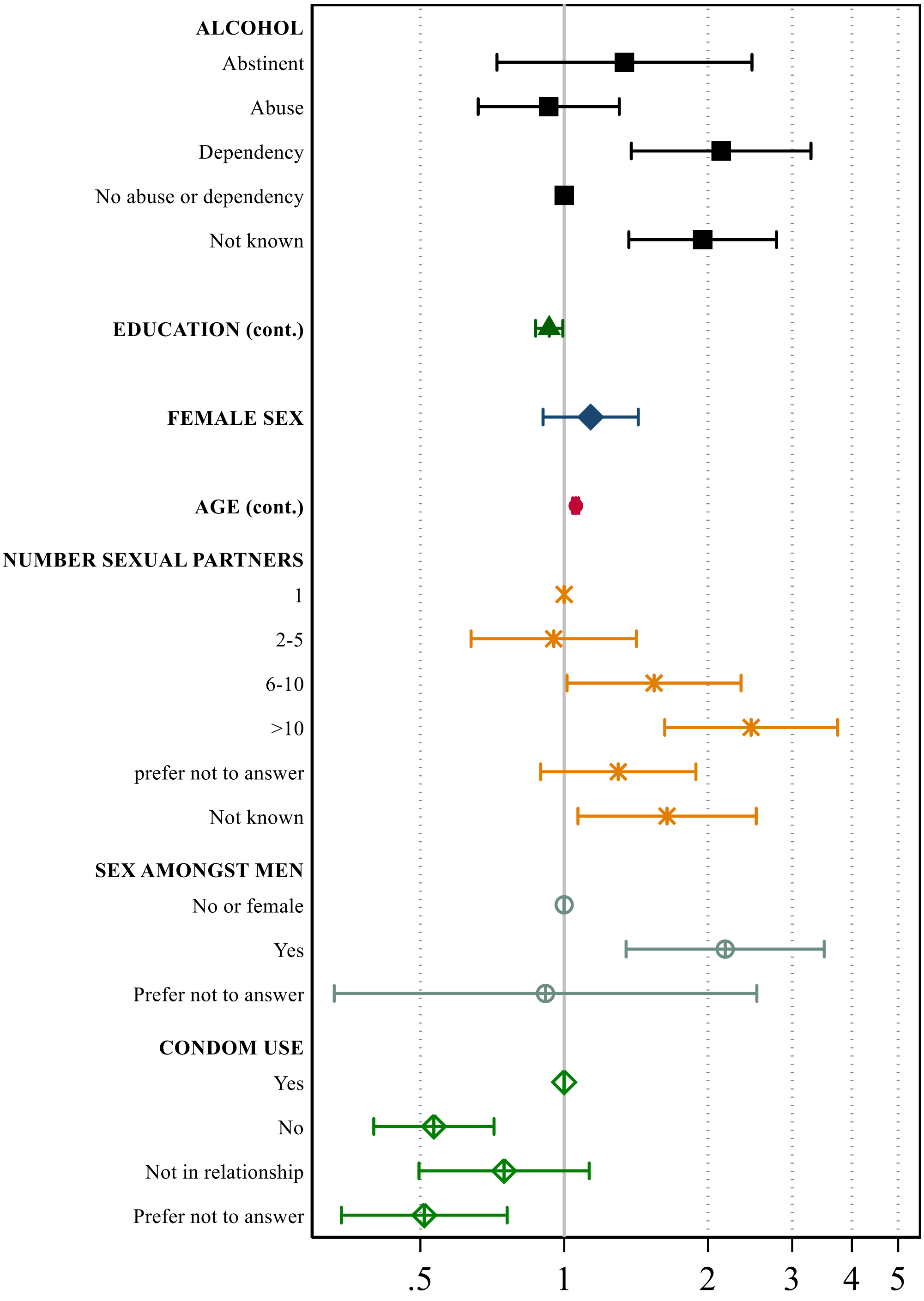

Supplementary Figure 4: ORs for Hepatitis B infection for covariates included in the fully adjusted models (adjusted for tattooed (yes/no)) for the screened/tested subsample.

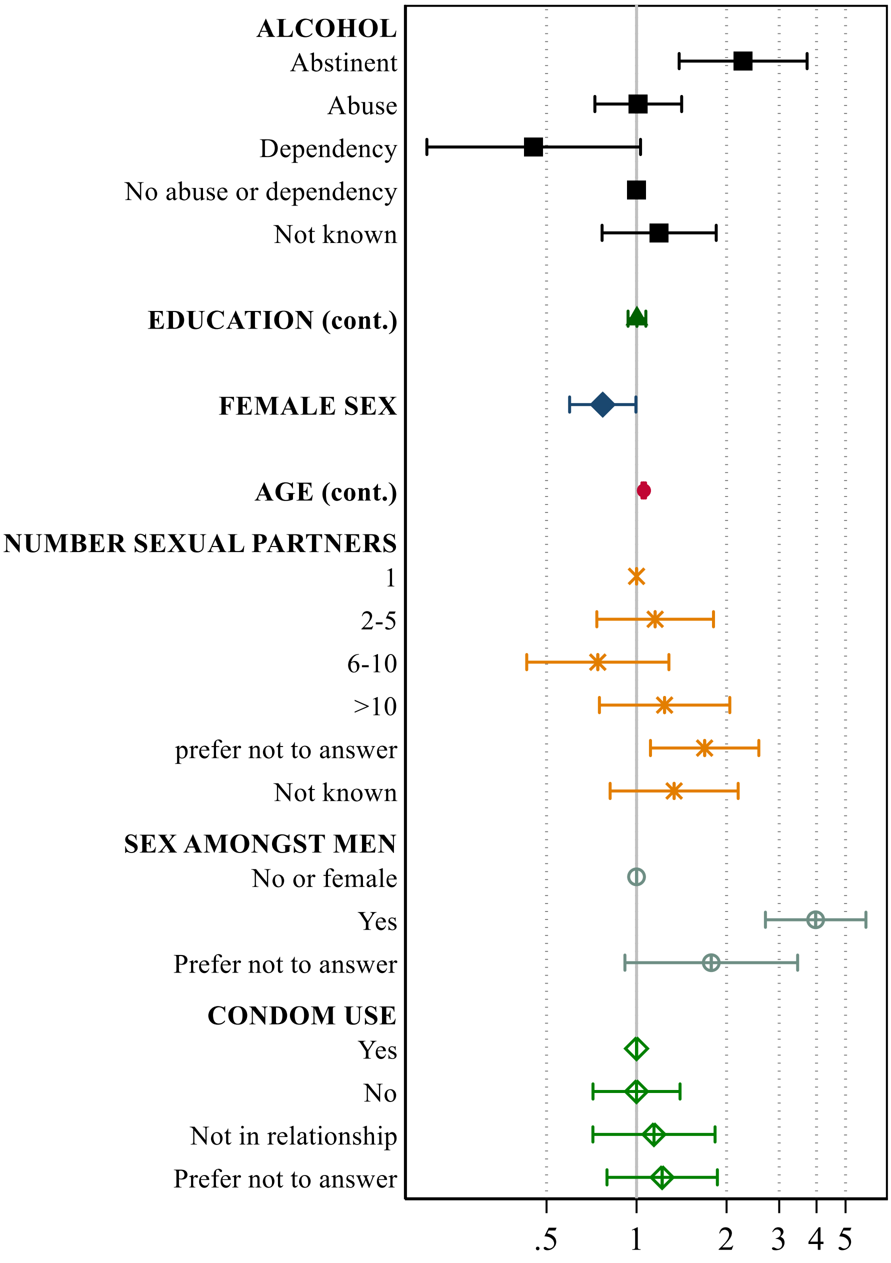

Supplementary Figure 5: ORs for Hepatitis C infection for covariates included in the fully adjusted models (adjusted for tattooed (yes/no)).for the screened/tested subsample.

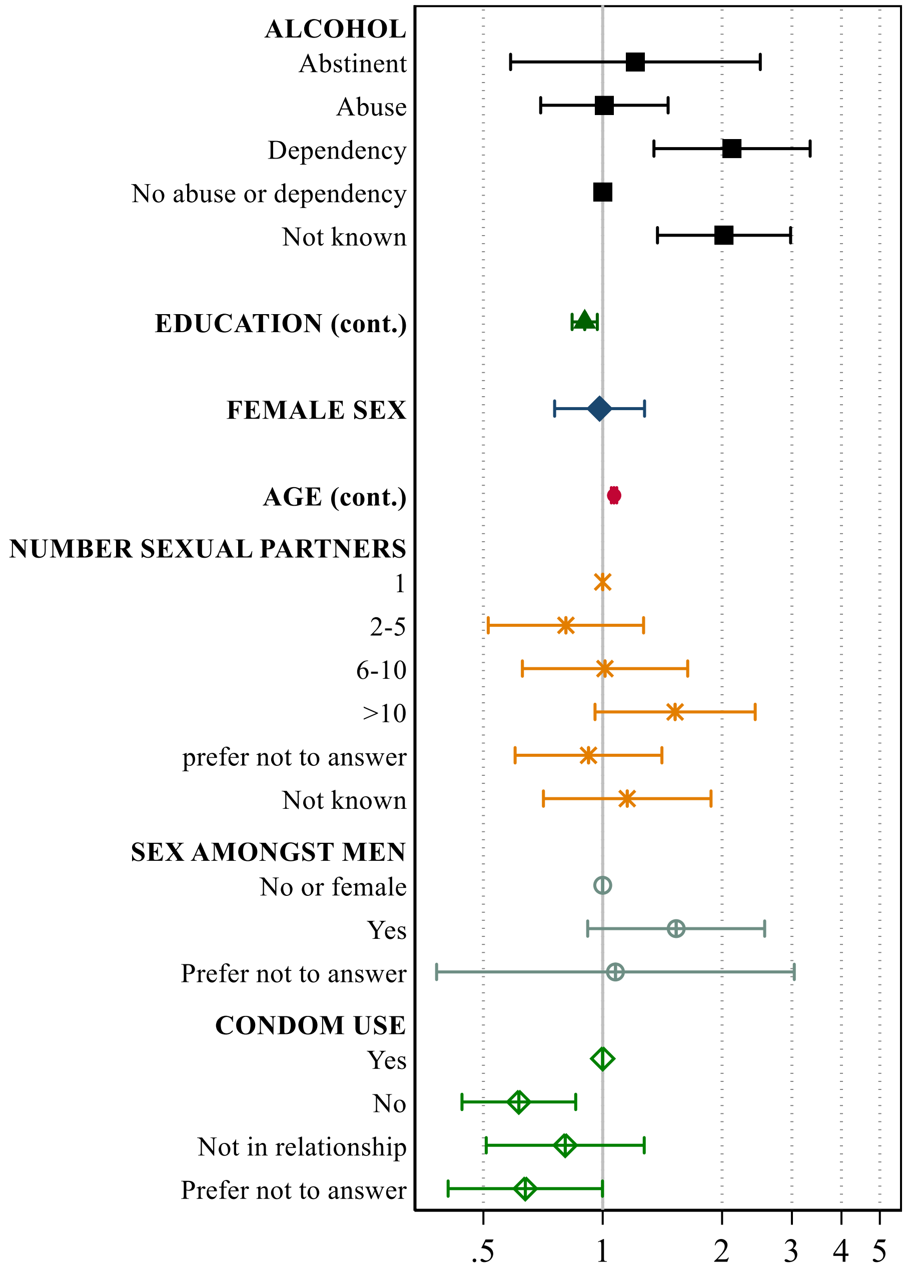

Supplementary table 1: Demographic characteristics and risk profile of participants ever infected with hepatitis B (HBV) or hepatitis C (HCV) infections or neither infection.

| | HBV 823 | HCV 476 | No infections[A] 109,620 | p value[B] |
|---|---|---|---|---|
| **Tattooed** | n (%) | n (%) | n (%) | |
| No | 746 (90.6 %) | 388 (81.5 %) | 96539 (88.4 %) | <0.001 |
| Yes | 77 (9.4 %) | 88 (18.5 %) | 12639 (11.6 %) | |
| **Sex** | | | | |
| Male | 422 (51.3 %) | 251 (52.7 %) | 49394 (45.2 %) | <0.001 |
| Female | 401 (48.7 %) | 225 (47.3 %) | 59784 (54.8 %) | |
| **Age** | | | | |
| <=45 | 46 (5.6 %) | 31 (6.5 %) | 32760 (30 %) | <0.001 |
| >45-<=55 | 99 (12 %) | 66 (13.9 %) | 24334 (22.3 %) | |
| >55-<=65 | 268 (32.6 %) | 198 (41.6 %) | 24834 (22.7 %) | |
| >65 | 410 (49.8 %) | 181 (38 %) | 27250 (25 %) | |
| **Education** | | | | |
| No diploma | 23 (2.8 %) | 21 (4.4 %) | 1749 (1.6 %) | <0.001 |
| General education certificate | 55 (6.7 %) | 41 (8.6 %) | 5160 (4.7 %) | |
| Vocational training certificate | 130 (15.8 %) | 76 (16 %) | 14780 (13.5 %) | |
| Baccalaureate or equivalent diploma | 113 (13.7 %) | 82 (17.2 %) | 16412 (15 %) | |
| Baccalaureate + 2 or 3 years | 207 (25.2 %) | 113 (23.7 %) | 29254 (26.8 %) | |
| Baccalaureate + 4 years | 68 (8.3 %) | 40 (8.4 %) | 10166 (9.3 %) | |
| Baccalaureate + 5 and more | 210 (25.5 %) | 87 (18.3 %) | 29689 (27.2 %) | |
| Missing | 17 (2.1 %) | 16 (3.4 %) | 1968 (1.8 %) | |
| **Alcohol consumption (audit score)** | | | | |
| Abstinent | 26 (5.6 %) | 20 (4.1 %) | 3291 (3 %) | <0.001 |
| Abuse | 65 (14.1 %) | 57 (11.6 %) | 17051 (15.5 %) | |
| Dependent | 18 (3.9 %) | 43 (8.8 %) | 4215 (3.8 %) | |
| Neither Abuse nor debendence | 312 (67.5 %) | 314 (64 %) | 78500 (71.6 %) | |
| Missing | 38 (8.2 %) | 53 (10.8 %) | 6255 (5.7 %) | |
| **Ever opioid replacement therapy** | 8 (1 %) | 19 (4 %) | 125 (0.1 %) | |
| **HIV positive** | 39 (4.7 %) | 33 (6.9 %) | 353 (0.3 %) | |
| **Number of sexual partners** | | | | |
| 1 partner | 100 (12.2 %) | 48 (10.1 %) | 17371 (15.9 %) | <0.001 |
| 2-5 partners | 143 (17.4 %) | 78 (16.4 %) | 30759 (28.2 %) | |
| 6-10 partners | 80 (9.7 %) | 63 (13.2 %) | 16108 (14.8 %) | |
| >10 partners | 106 (12.9 %) | 91 (19.1 %) | 10861 (9.9 %) | |
| Prefer not to answer | 267 (32.4 %) | 117 (24.6 %) | 22818 (20.9 %) | |
| Missing | 127 (15.4 %) | 79 (16.6 %) | 11261 (10.3 %) | |
| **Sex amongst men** | | | | |
| Women or never | 291 (69 %) | 200 (79.7 %) | 45169 (91.4 %) | <0.001 |
| Ever | 110 (26.1 %) | 45 (17.9 %) | 3056 (6.2 %) | |
| Prefer not to answer | 21 (5 %) | 6 (2.4 %) | 1169 (2.4 %) | |
| **Current condom use** | | | | |
| Yes | 144 (17.5 %) | 106 (22.3 %) | 27375 (25.1 %) | <0.001 |
| No | 443 (53.8 %) | 229 (48.1 %) | 58887 (53.9 %) | |
| Not currently in a relationship | 80 (9.7 %) | 56 (11.8 %) | 8722 (8 %) | |
| Missing | 156 (19 %) | 85 (17.9 %) | 14194 (13 %) | |

[A] Including 55 participants with cHBV/cHCV coinfections

[B] p-value calculated for no infection against any infection

Supplementary Table 2: Odd's ratios and 95% CIs of the multivariate logistic regression models on risk of chronic Hepatitis B or chronic Hepatitis C virus infection and tattoo exposure variables for subgroup analyses of the screened/tested population and a subgroup with low risk profile (excluding participants with alcohol abuse, having slept >10 partners, or having had sex amongst men).

| | Screened/tested sample[A] | | | | | | Low-risk sample[B] | | | | | |
|---|---|---|---|---|---|---|---|---|---|---|---|---|
| | Hepatitis | | HBV | | cHCV | | Hepatitis | | HBV | | cHCV | |
| | OR | 95% CI | OR | 95% CI | OR | 95% CI | OR | 95% CI | OR | 95% CI | OR | 95% CI |
| **Tattooed** | 1.47 | (1.12; 1.93) | 1.06 | (0.69; 1.62) | 2.1 | (1.48; 2.98) | 1.25 | (0.86; 1.81) | 1.12 | (0.68; 1.82) | 1.58 | (0.94; 2.66) |
| **Tattoo context** | | | | | | | | | | | | |
| Not tattooed | | ref | | ref | | ref | | ref | | ref | | ref |
| Only in studio | 0.5 | (0.26; 0.93) | 0.54 | (0.24; 1.22) | 0.57 | (0.23; 1.41) | 0.34 | (0.2; 1.44) | 0.27 | (0.07; 1.08) | 0.61 | (0.19; 1.93) |
| At least one tattoo outside parlour | 2.64 | (1.56; 4.47) | 1.22 | (0.44; 3.35) | 4.25 | (2.3; 7.86) | 2.79 | (2.26; 7.94) | 2.69 | (1.18; 6.12) | 3.75 | (1.63; 8.62) |
| **Tattoo country** | | | | | | | | | | | | |
| Not tattooed | | ref | | ref | | ref | | ref | | ref | | ref |
| EU/EFTA countries with regulation | 0.91 | (0.58; 1.43) | 0.56 | (0.26; 1.19) | 1.38 | (0.79; 2.42) | 0.78 | (0.42; 1.43) | 0.68 | (0.3; 1.53) | 1.18 | (0.55; 2.54) |
| No or recent regulation | 1.46 | (0.59; 3.63) | 1.53 | (0.47; 4.96) | 2.02 | (0.63; 6.48) | 2.08 | (0.66; 6.56) | 2.24 | (0.55; 9.13) | 3.24 | (0.79; 13.25) |
| **Interaction amongst the tattooed** | | | | | | | | | | | | |
| In studio in regulating country | | ref | | ref | | ref | | ref | | too few obs/cell | | too few obs/cell |
| Outside parlour in regulating country | 7.22 | (2.56; 20.38) | 6.2 | (0.68; 56.2) | 7.76 | (2.18; 27.64) | 10.4 | (2.55; 42.46) | | | 10.26 | (1.71; 61.68) |
| Inside parlour outside in regulating country | 2.36 | (0.53; 10.55) | 7.02 | (0.72; 68.36) | 0.9 | (0.08; 9.96) | 3.03 | (0.29; 31.32) | | | 4.42 | (0.35; 55.66) |
| Outside parlour outside in regulating country | 8.98 | (2.22; 36.36) | 18.78 | (1.67; 210.57) | 15.39 | (2.6; 91.14) | 12.38 | (1.87; 82.14) | | | 7.71 | (0.58; 102.2) |

[A]Excluding HIV positive participants and ever injected drug users; adjusted for sex, age at initial tattoo assessment, highest educational level, alcohol consumption (audit score), number of sexual partners, condom use, and sex amongst men

[B]Excluding HIV positive participants , ever injected drug users, participants with known alcohol abuse, those who slept with >10 partners, and having had sex amongst men; adjusted for sex, age at initial tattoo assessment, highest educational level, alcohol consumption (audit score), number of sexual partners, condom use

#### Supplementary table 3: Results of sensitivity analyses stratified by biological sex.

| | Hepatitis | | | | cHBV | | | | cHCV | | | |
|---|---|---|---|---|---|---|---|---|---|---|---|---|
| | Female | | Male | | Female | | Male | | Female | | Male | |
| | ORA | 95% CI | ORA | 95% CI | ORA | 95% CI | ORA | 95% CI | ORA | 95% CI | ORA | 95% CI |
| **Tattooed** | 1.18 | (0.83; 1.68) | 1.61 | (1.19; 2.19) | 0.73 | (0.42; 1.27) | 1.23 | (0.82; 1.86) | 1.97 | (1.26; 3.11) | 2.37 | (1.56; 3.6) |
| **Tattoo context** | | | | | | | | | | | | |
| Not tattooed | ref | | ref | | 0.46 | | ref | | ref | | ref | |
| Only in studio | 0.32 | (0.13; 0.77) | 0.52 | (0.16; 1.66) | 0.31 | (0.1; 0.97) | 0.6 | (0.25; 1.48) | 0.46 | (0.15; 1.48) | 0.61 | (0.19; 1.92) |
| At least one tattoo outside parlour | 2.9 | (1.47; 5.71) | 1.21 | (0.29; 4.96) | 1.53 | (0.48; 4.83) | 1.87 | (0.86; 4.03) | 5.37 | (2.44; 11.84) | 3.67 | (1.83; 7.37) |
| **Tattoo country** | | | | | | | | | | | | |
| Not tattooed | ref | | ref | | ref | | ref | | ref | | ref | |
| EU/EFTA countries with regulation | 0.77 | (0.44; 1.35) | 0.89 | (0.53; 1.51) | 0.47 | (0.19; 1.16) | 0.77 | (0.38; 1.57) | 1.3 | (0.65; 2.61) | 1.38 | (0.69; 2.75) |
| No or recent regulation | 0.49 | (0.07; 3.52) | 2.47 | (1.07; 5.7) | 0.83 | (0.11; 5.94) | 2.48 | (0.89; 6.94) | 1.11 | (0.15; 8.09) | 3.3 | (1.03; 10.58) |
| **Interaction amongst the tattooed** | | | | | | | | | | | | |
| In studio in regulating country | | | ref | | | | ref | | | | ref | |
| Outside parlour in regulating country | too few obs | | 3.17 | (0.93; 10.86) | too few obs | | 3.39 | (0.62; 18.53) | too few obs | | 4.45 | (0.81; 24.54) |
| Inside parlour outside in regulating country | | | 4.48 | (0.95; 21.1) | | | 4.81 | (0.64; 36.38) | | | 4.38 | (0.34; 56.24) |
| Outside parlour outside in regulating country | | | 10.67 | (2.7; 42.13) | | | 14.98 | (2.42; 92.87) | | | 14.56 | (2.12; 100.2) |

[A]Adjusted for sex, age at initial tattoo assessment, highest educational level, alcohol consumption (audit score), number of sexual partners, condom use, and sex amongst men (for men only)

Supplementary Table 4: Results of sensitivity analysis when restricting to the period after 31st December 2006 for which timely order of tattooing before/after infection is known.

| | HBV/HCV | | HBV | | HCV | |
|---|---|---|---|---|---|---|
| | OR[A] | 95% CI | OR[A] | 95% CI | OR[A] | 95% CI |
| **Tattooed** | 1.5 | (0.95; 2.36) | 0.96 | (0.45; 2.02) | 2.14 | (1.28; 3.58) |
| **Tattoo context** | | | | | | |
| not tattooed | | ref | | ref | | ref |
| only in studio | 0.44 | (0.14; 1.39) | 0.56 | (0.14; 2.29) | 0.25 | (0.04; 1.82) |
| outside studio | 3.75 | (1.81; 7.78) | 1.64 | (0.4; 6.77) | 6.35 | (3.01; 13.39) |
| **Tattoo country** | | | | | | |
| Not tattooed | | ref | | | | ref |
| France only | 1.25 | (0.65; 2.4) | too small n per cell | | 1.72 | (0.82; 3.58) |
| At least one tattoo outside France | 0.99 | (0.14; 7.13) | | | 1.68 | (0.23; 12.17) |

[A]Adjusted for sex, age at initial tattoo assessment 2020
